# Nasopharyngeal SARS-CoV2 viral loads in young children do not differ significantly from those in older children and adults

**DOI:** 10.1101/2020.09.17.20192245

**Authors:** Sharline Madera, Emily Crawford, Charles Langelier, Nam K Tran, Ed Thornborrow, Steve Miller, Joseph L DeRisi

**Affiliations:** Department of Medicine, Division of Infectious Diseases University of California, San Francisco, CA; Chan-Zuckerberg Biohub, San Francisco, CA; Department of Microbiology and Immunology University of California San Francisco, CA; Department of Medicine and Biochemistry & Biophysics University of California San Francisco, CA; Department of Pathology and Laboratory Medicine University of California, Davis School of Medicine, Sacramento, CA; Department of Laboratory Medicine University of California San Francisco, CA

## Abstract

The role of children in the spread of the SARS-CoV2 coronavirus has become a matter of urgent debate as societies in the US and abroad consider how to safely reopen schools. Small studies have suggested higher viral loads in young children. Here we present a multicenter investigation on over five thousand SARS-CoV-2 cases confirmed by real-time reverse transcription (RT) PCR assay. Notably, we found no discernable difference in amount of viral nucleic acid among young children and adults.

## Main

Children remain underrepresented in current studies aimed to analyze the spread of SARS-CoV2 coronavirus, making their contribution to viral transmission elusive. It is well established that, in general, children experience less severe illness than do adults, though in rare cases children can be subject to a severe multisystem inflammatory syndrome (1). And there is an emerging view that children may play a lesser role in the spread of Coronavirus Disease 2019 (COVID-19) than they do in other respiratory illnesses (2), but much uncertainty about this remains (3). Recently, it was reported that children less than five years old may carry higher viral loads in the nasopharynx than older children and adults (4), raising concerns that exposure to this group may pose special epidemiologic risks. Here we report results bearing on this question from two coronavirus testing laboratories that serve large populations of patients in California.

Laboratory A serves the UC San Francisco health care system, as well as local clinics and also provides tests to the county health departments in 26 California counties. Laboratory B serves principally the UC Davis health care system and partner clinics/hospitals centered in Sacramento, CA. Both labs test similarly-collected nasopharyngeal swab specimens. Both laboratories employed real-time RT-PCR SARS-CoV2 assays, and record for each positive sample the cycle threshold (Ct) value. In general, primers from both the E and N genes are employed, though in this analysis for simplicity we show only the E gene Ct values. For both labs, we report the results of all specimens tested from March-August 2020. The population is predominantly comprised of out-patients representing both symptomatic clinical cases and asymptomatic cases identified by contact tracing.

Among 5,544 patients with laboratory-confirmed COVID-19 spanning Laboratory A and B, 199 children aged less than five years, 665 youth aged five to seventeen, and 4680 adults aged eighteen and older, were identified. In order to ascertain potential differences in viral load carriage, Ct values were queried across age groups. Ct values were used to approximately reflect the viral load (inversely related to Ct value). As depicted in Figure 1, no significant differences in Ct value were observed across the three groups. In particular, the children less than age 5 did not display higher nasopharyngeal viral loads than older children or adults.

**Figure 1:**
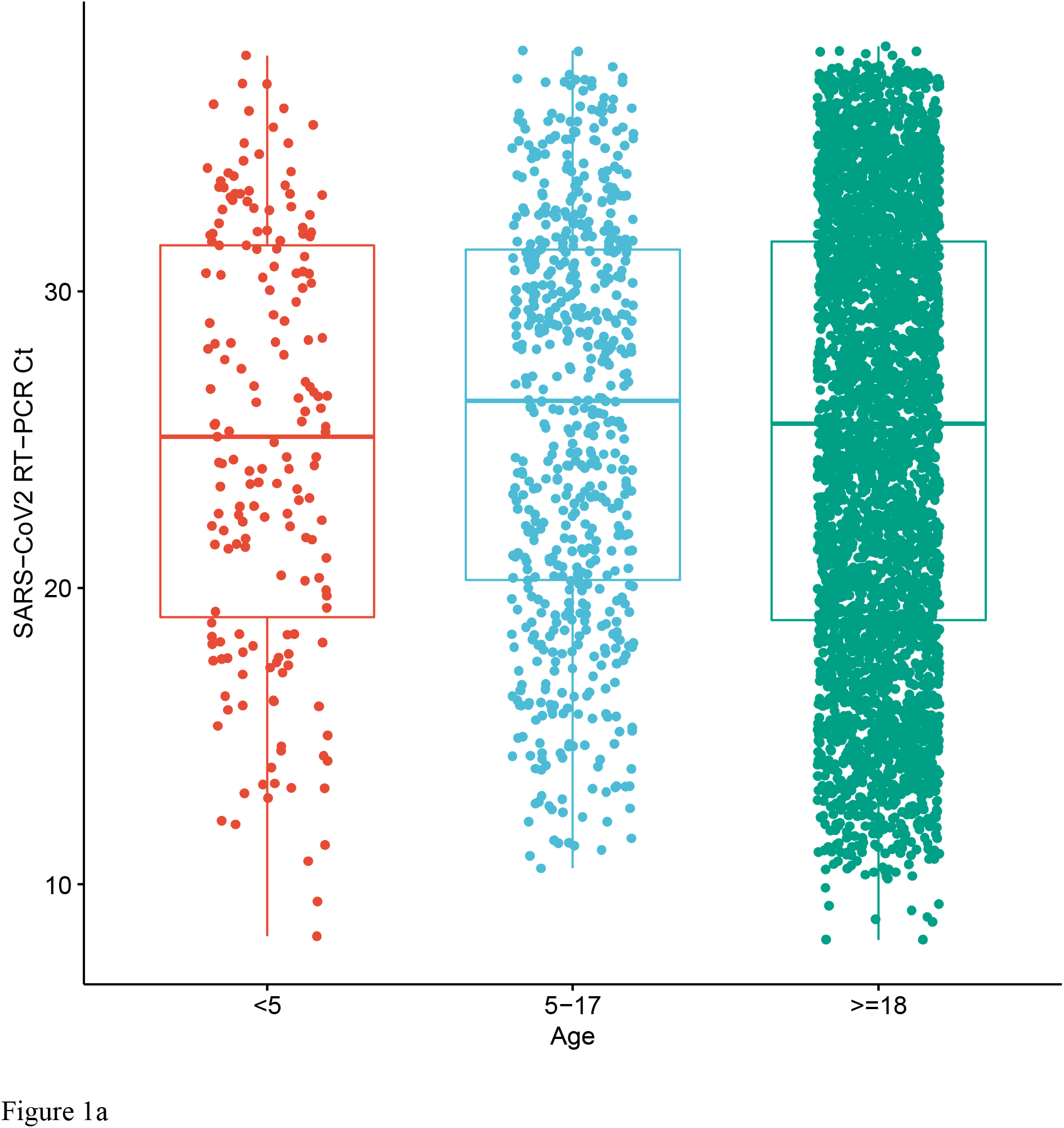
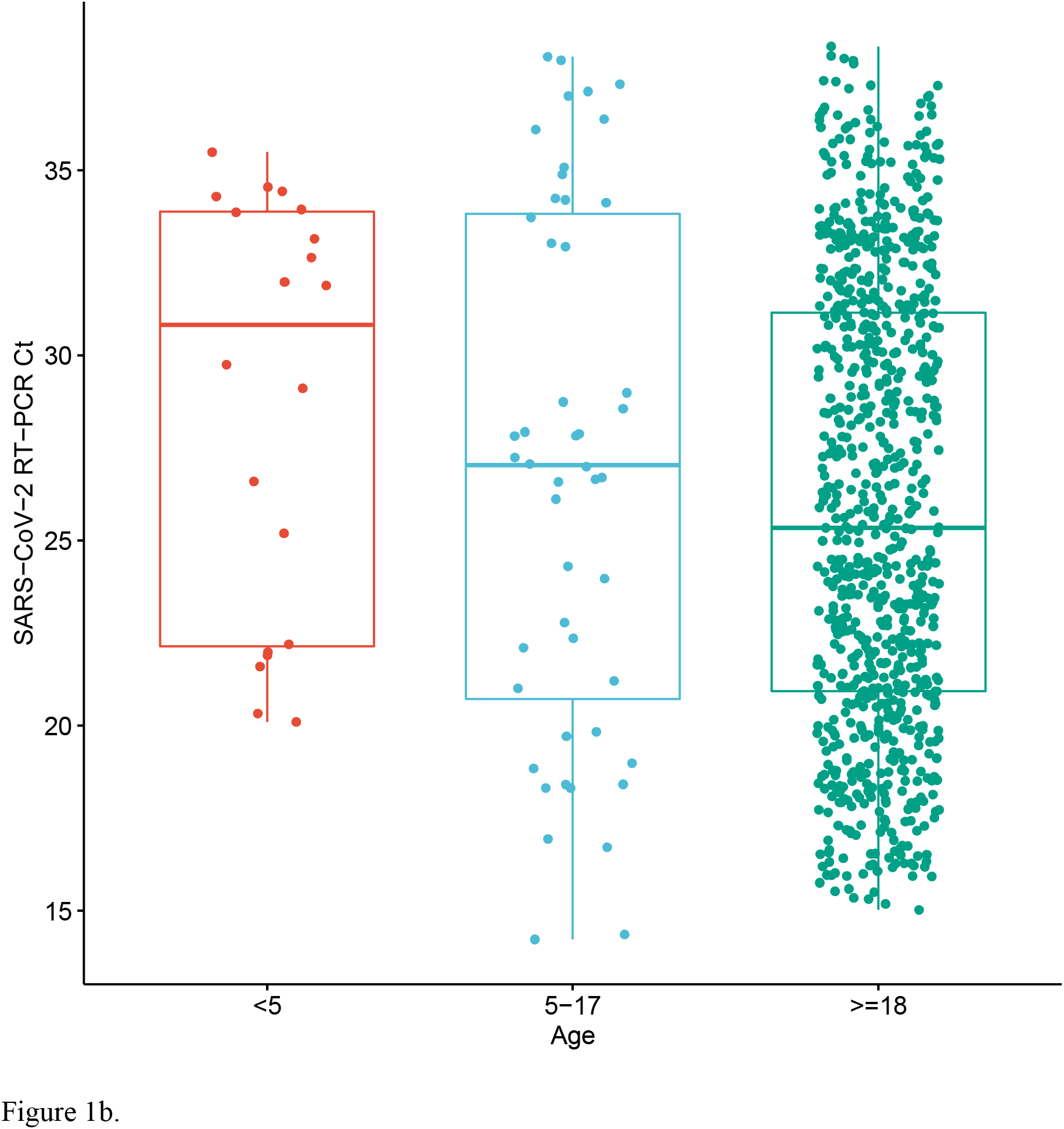
Age distributed nasopharyngeal SARS-CoV2 viral nucleic acid content. SARS-CoV2 viral nucleic acid detected by real-time RT-PCR in nasopharyngeal swabs from patients infected with SARS-CoV-2 as detected by (a) laboratory A (N = 4,619, ANOVA p = 0.18) and (b) laboratory B (N = 925, ANOVA p = 0.073). Data are stratified by three age groups, ages < 5; 5–17; 18 and older.

In contrast to a prior smaller study (4), we do not observe higher nasopharyngeal viral loads in children below the age of 5 years. The population studied is largely reflective of asymptomatic and mildly symptomatic individuals, and notably does not include patients hospitalized with severe disease. Thusly, these conclusions apply primarily to the outpatient setting and may not extend to the severely-ill. There are conflicting data on the association of viral load with disease severity, with some studies showing higher viral loads in severe cases (5), while others indicate a lower viral load in hospitalized patients than those not hospitalized (6). Since the viral load changes rapidly during early infection, the time between symptom onset and sampling is a significant variable. Some patients with severe disease may delay entry to care, missing the peak period of viral shedding. Notably, these earlier studies were done primarily in the hospital setting, so the findings do not necessarily translate to the outpatient population. Nevertheless, the robust sample size of 5,544 patients presented in this work, including close to 200 children aged less than 5 years, is more likely to be representative of the general population of infected subjects. An accurate understanding of the variables that affect viral transmission, including amount of virus carriage, will be essential to guide public policy efforts as re-opening strategies are devised.

We caution, however, that viral load as determined by RT-PCR is only one of many potential influences on infectivity. PCR accurately enumerates viral genomes, but does not indicate whether they come from infectious virions, defective viral particles, or lysed infected cells. Infectivity in populations is affected by many other clinical, behavioral and environmental factors. Our findings argue against the idea that young children are more infectious due to higher viral loads, and suggest an alternative explanation for their contribution to SARS-CoV2 transmission, such as representing a reservoir of asymptomatic infections. Ultimately, the role of young children in the spread of SARS-CoV2 can only be determined by careful clinical and epidemiological studies that examine transmission events in the population.

## Method

### Real time RT–PCR assay for SARS-CoV-2 RNA

Nasopharyngeal swabs collected into DNA/RNA Shield (Laboratory A) or viral transport media (Laboratory B) were obtained from patients. At Laboratory A, samples were subjected to a clinically reportable Laboratory Developed Test (LDT) consisting of total nucleic acid extraction using the Zymo Quick-DNA/RNA Viral Magbead kit (Zymo Research, Orange CA) and real-time RT–PCR using the NEB Luna Universal RT-qPCR kit (New England Biolabs, Ipswitch MA) on Bio-Rad CFX384 instruments (Bio-Rad, Hercules, CA). At Laboratory B, automated nucleic extraction and real-time RT-PCR testing was performed on the Roche cobas 6800 (Roche Diagnostics, Indianapolis, IN) using the EUA SARS-CoV-2 assay. Each real-time RT– PCR assay provided a threshold cycle (Ct) value, indicating the number of cycles surpassing the threshold for a positive test. Samples were considered positive if the Ct value was ≤ 40, and otherwise it was negative.

### Statistics

A one-way analysis of variance was used to compare Ct values across age groups of samples positive for COVID-19 infection with R ggpubr v.0.4.0.

## Data Availability

The data shown in the manuscript are available upon request from the corresponding author.

## Acknowledgements

This work was funded by the Chan Zuckerberg Biohub. We greatly appreciate the guidance Don Ganem in the preparation of this manuscript.

## Author information

### Contributions

E.C., E.T., J.L.D., N.T. contributed to study design. S. Madera, E.C., J.L.D., and N.T. contributed to collection of data. S. Madera contributed to data analysis. S. Madera, E.C., N.T., J.L.D., and S. Miller contributed to manuscript preparation. All authors approved the manuscript.

## Ethics Declaration

### Competing Interests

Nam Tran is a consultant for Roche Diagnostics. UC Davis Health is a designated Roche Molecular Center of Excellence.

## Notes

### Author Declarations

IRB Oversight body: University of California, San Francisco Institutional Review Board. Per the UCSF IRB review board guidelines, we have provided a certification form from our IRB/oversight body denoting that our research did not require IRB review at UCSF. The form is attached under "Files Metadata"

